# Clinical and genetic insights of Parkinson’s Disease in a Mexican cohort: highlighting Latino’s diversity

**DOI:** 10.1101/2023.08.28.23294700

**Authors:** Alejandra Lázaro-Figueroa, Paula Reyes-Pérez, Eugenia Morelos-Figaredo, Carlos M. Guerra-Galicia, Ingrid Estrada-Bellmann, Karla Salinas-Barboza, Yamil Matuk-Pérez, Nadia A. Gandarilla-Martínez, Dante Oropeza, Ulises Caballero-Sánchez, Pablo Montés-Alcántara, Araliz López-Pintor, Ana P. Angulo-Arrieta, Victor Flores-Ocampo, Ian M. Espinosa-Méndez, Alejandra Zayas-Del Moral, Edith Gaspar-Martínez, Damaris Vazquez-Guevara, Mayela Rodríguez-Violante, Emily Waldo, Thiago P. Leal, Miguel Inca-Martinez, Ignacio F. Mata, Sarael Alcauter, Miguel E. Rentería, Alejandra Medina-Rivera, Alejandra E. Ruiz-Contreras, the Mexican Parkinson’s Research Network (MEX-PD) and the Latin American Research Consortium on the Genetics of Parkinson’s Disease (LARGE-PD)

**Author notes:** Corresponding authors Sarael Alcauter, Instituto de Neurobiología, Universidad Nacional Autónoma de México, Alejandra Medina-Rivera, Laboratorio Internacional de Investigación sobre el Genoma Humano, Universidad Nacional Autónoma de México, Alejandra E. Ruiz-Contreras, Laboratorio de Neurogenómica Cognitiva, Facultad de Psicología Facultad de Psicología, Universidad Nacional Autónoma de México. These authors contributed equally to this work (Alejandra Lázaro-Figueroa, Paula Reyes-Pérez). These authors jointly led and supervised the research.

## Abstract

**Objective:** This study aims to describe the establishment of the Mexican Parkinson’s Research Network (MEX-PD), a consortium dedicated to investigating the clinical, genetic, environmental, and neurophysiological underpinnings of phenotypic diversity in Mexican Parkinson’s disease (PD) patients and to present preliminary clinical and genetic outcomes.

**Methods:** PD patients and control participants were recruited from medical centers across Mexico. The initial recruitment phase involved comprehensive neurological evaluations, cognitive assessments, and DNA collection. We conducted classical statistical analyses on clinical variables. Secondly, following genotyping with NeuroBooster array, quality control and imputation, preliminary analysis of ancestry composition, allele frequency calculation and association analysis was carried out for 294 samples.

**Results:** The cohort consisted of 530 control participants and 470 PD patients, with a mean age of diagnosis of 59.9 ± 11.52 years. Among the PD patients, 21.2% were identified as having early-onset PD (<50 years old). Ancestry composition analysis revealed that the main components were European (49.8% in cases and 42.4% in controls) and Native American (46.1% in cases and 54.3% in controls). Variants in genes such as *NOTCH*, *LRRK2*, *MTHFR* and *KPNA1* emerged as relevant to be prioritized in future studies.

**Conclusions:** The MEX-PD consortium will contribute to the understanding of PD within the Mexican population. The data collected will enable a deeper comprehension of the specific contributions of genetic and environmental factors to these outcomes.

**Significance:** This research advocates for the development of personalized treatments and aims to improve the quality of life for Mexican PD patients.

## 1. Introduction

Parkinson’s Disease (PD) is the second most prevalent neurodegenerative disease worldwide (Tolosa et al., 2021), affecting more than 100 people per 100,000 and summing to over 8.51 million affected people worldwide (Ou et al., 2021). This represents a 155.5% increase in cases from 1990 to 2019. Due to the increase in life expectancy, the prevalence of PD is expected to rise as well (Dorsey et al., 2018). This disease is associated with the loss of dopaminergic neurons in the *substantia nigra* of the brainstem. PD pathology has also been linked to the intraneuronal accumulation of α-synuclein proteins (Aarsland et al., 2021; Bloem et al., 2021).

Given its motor symptoms, such as resting tremor, rigidity, postural instability, and bradykinesia, PD is classified as a movement disorder (Bloem et al., 2021). However, it also shows non-motor symptomatology, including hyposmia, apathy, anxiety, depression, sleep disorders (e.g., insomnia or daytime sleepiness), mild cognitive impairment (MCI), and dementia (Goldman et al., 2018; Ben-Shlomo et al., 2024). The appearance of these symptoms in PD varies in presence and severity between patients, which contributes to the heterogeneity of the disease. (Kalia & Lang, 2015).

Despite clinical efforts at major health centers in Mexico’s largest cities and ongoing biomedical research, there remains a significant need for increased research into the genetic and environmental risk factors for PD in the Mexican population. To our knowledge, no national epidemiological PD study exists in Mexico to help identify these risk factors, mainly due to the economic and logistic complexity of such an effort. The incidence of PD in Mexico is estimated to be 9.48 per 100,000 person-years for individuals aged over 20 years old (Rodríguez-Violante et al., 2011; Rodríguez-Violante et al., 2019). It was anticipated that by 2023, this incidence will rise to 14.9 per 100,000 (Martínez-Ramírez et al. 2020), but no study to date has confirmed this projection. The highest incidence rates are reported in the states of Sinaloa (27.6 per 100,000), Colima (23.5 per 100,000), and Durango (20 per 100,000; Rodríguez-Violante et al., 2011; Martínez-Ramírez et al., 2020). The hypothesis to explain the difference in incidence is that, in these high-prevalence states, the population is more exposed to environmental factors, such as pesticides or contaminated water, related to their agricultural economy. Thus, the disparity between the prevalence of PD in Mexico compared with that worldwide indicates that genetic or/and environmental factors might account for it.

Furthermore, while there are successful efforts to obtain genetic resources for demographic and evolutionary studies in Mexico, such as the Mexico City Prospective Study (Ziyatdinov et al. 2023), and the Mexican Biobank (Sohail et al. 2023) these projects collect limited information on family history, mental health, cognition, and chronic and degenerative diseases, and they do not have specific information about PD.

Additionally, to our knowledge, most genetic PD studies in Mexicans have relied on targeted genotyping or sequencing of selected variants in known PD risk genes. Despite this limitation, findings highlight the significant role of genetic diversity and admixture in Latino populations. For instance, a study identified the rs1491942 variant in the *LRRK2* gene as a risk factor for PD (OR=2.26, p=0.01), especially among those with a high proportion of Native American ancestry (≥56.6%; Romero-Gutiérrez et al., 2021). Additionally, the rs1801133 variant in the *MTHFR* gene was more common in PD patients with 32-52% Native American ancestry compared to controls (OR=2.02, p=0.043; Romero-Gutiérrez et al., 2021).

Nevertheless, Mexico has yet to conduct a genome-wide association study (GWAS) or similar large-scale genetic research on PD. Hence, there is a pressing need to understand how genetics contribute to PD and interact with environmental factors in highly admixed populations. The Mexican population, which exhibits three-way admixture deriving from Native American (37-49%), European (50-60%), and African (1-3%) ancestries (Cerda-Flores et al., 2002; Moreno-Estrada et al., 2014), presents a unique opportunity for this study.

In response to this need, the Mexican Parkinson’s Research Network (MEX-PD; Red Mexicana de Investigación en Parkinson, in Spanish) was established in 2021. This national research consortium brings together experts in genetics, human cognition, functional neuroimaging, and neurologists specialized in movement disorders. This network aims to investigate the prevalence, clinical and cognitive features, comorbidities, and the evolution of PD in the Mexican population and to identify diagnostic and progression biomarkers based on cognitive, genetic and neuroimaging evaluations. Additionally, using a whole-genome screening approach will enable a better understanding of the genetic underpinnings of PD in Mexico, considering ancestry-specific factors. Our findings may have implications beyond Mexico, providing valuable insights for comparable groups, such as other Latin American countries and Latinos in the United States of America and other countries (Aamodt et al., 2023).

The goal of this manuscript is two-fold: First, to present cohort-based information about MEX-PD and the phenotypical and genotypical information we are collecting, aiming to promote local collaborations to increase the cohort, as well as international collaborations and show preliminary clinical outcomes; Second, to analyze the ancestry of our cohort and the allele frequency of genetic variants, for comparing it with that from European and African populations.

## 2. Methods

### 2.1 Participants

At the time of writing this manuscript, June of 2024, our study has enrolled 530 healthy control participants and 470 PD patients. PD diagnosis is confirmed by neurologists specialized in movement disorders, based on the UK Brain Bank Criteria. The inclusion criteria for patients are as follows: a) diagnosis of idiopathic PD; b) born in Mexico; c) majority of lifetime spent in Mexico, at least the half of their lives; d) provision of written informed consent; e) at least 18 years of age. The same criteria, excluding a PD diagnosis are applied for the control group, supplemented by: f) age at examination of at least 45 years old; g) no consanguineous relationship with individuals with PD; and h) no diagnosis of any neurodegenerative disease. Controls are recruited from medical centers (e.g., spouses or caregivers of PD patients without a genetic relationship), public events (such as musical shows and science fairs), and parks. Patients with PD have been recruited through both public and private medical centers in twenty Mexican states (Figure 1). All participants are requested to donate a DNA sample via buccal swab for subsequent genotyping.

**Figure 1.**
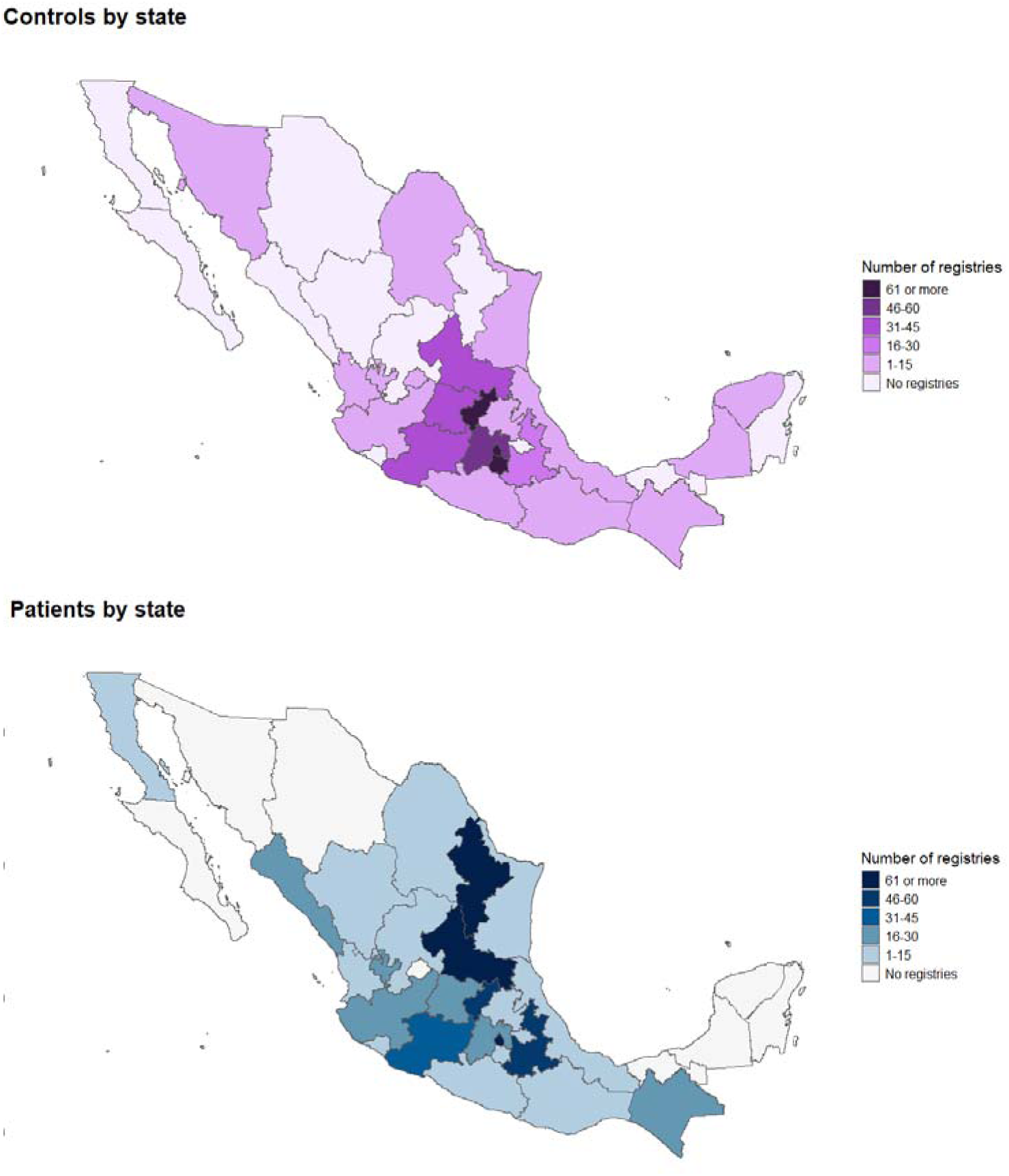
Maps of Mexico with the geographical distribution of participants by state.

Institutional support and the necessary infrastructure are provided by the *Universidad Nacional Autónoma de México* (UNAM, may translate as National Autonomous University of Mexico). This research has received approval from the Bioethics Committee of the Institute of Neurobiology at UNAM. All participants sign the approved informed consent after an explanation of the research protocol and answer any questions they may have. Additionally, MEX-PD closely collaborates with the Latin American Research Consortium on the GEnetics of Parkinson’s Disease (LARGE-PD; Zabetian & Mata, 2017) which aims to enhance our understanding of genetics of PD in Latin America. LARGE-PD is coordinated from Cleveland Clinic located in Cleveland, Ohio, United States of America, more else, with its coordination, the collected data will be shared through the ASAP-GP2 community (GP2; 2021).

### 2.2 Data collection

Data collection is conducted online using the Research Electronic Data Capture (REDCap) platform (Harvey, 2018; Wright, 2016). REDCap is a secure platform where all survey data are recorded and locally saved on a database server at the National Laboratory of Advanced Scientific Visualization at UNAM; under stringent security protocols. Furthermore, sensitive data is pseudonymized for additional protection.

### 2.3 Instruments

Participants respond to an array of questionnaires, scales, psychological instruments, and neuropsychological batteries, all in Spanish language.

Data collection from participants is divided into three sessions. First, they complete a comprehensive record of their clinical history, anthropometric information, an environmental exposure questionnaire, and complete the Spanish version (Aguilar-Navarro et al., 2018) of the Montreal Cognitive Assessment (MoCA; Nasreddine et al., 2005), validated in the Mexican population. A neurologist applies the clinical history for PD patients, the official Movement Disorder Society–Unified Parkinson’s Disease Rating Scale (MDS-UPDRS; Goetz et al., 2008), the Hoehn-Yahr scale (Hoehn & Yahr, 2001), and records PD-related symptoms and medication prescriptions. Also, at this first session the buccal swabs were collected from participants to obtain DNA, so it was done at the site of the register.

Following the initial data collection session, participants are contacted via telephone. In this second session, they complete a series of instruments related to mental health, in Spanish: the State-Trait Anxiety Inventory (STAI; Díaz-Guerrero & Spielberger, 1975), State-Trait Depression Inventory (ST-DEP; Martín-Carbonell et al., 2012), Parkinson Anxiety Scale (Leentjens et al., 2014) and Symptom Checklist-90-R (SCL-90-R; Derogatis et al., 1976). STAI, SR-DEP and SCL-90-R are validated for the Mexican Population (Díaz-Guerrero & Spielberger, 1975, Martín-Carbonell et al., 2012, Cruz-Fuentes et al, 2005). The Cognitive Reserve Index Questionnaire (Nucci et al., 2012), measuring cognitive reserve is also included (Spanish version).

In the third and final session, participants are contacted via video call. This session aims to evaluate cognitive function using a computerized neuropsychological battery in Spanish called *Creyos* (initially known as the Cambridge Brain Sciences; https://creyos.com/features/tasks).

A description of all available questionnaires is provided in Supplementary Table 1.

### 2.4 Statistical analysis

Descriptive variables, such as age, sex, years of education, occurrence of head injury, and ancestry, were compared between control and PD patient groups. The Chi-squared test was employed to determine differences in sex and the presence of a head injury or concussion. Similarly, the Student’s t-test for independent samples was used to evaluate differences between groups in terms of age and years of education.

The clinical data was analyzed as follows. The mean and standard deviation [SD] of age of onset and disease duration (calculated from the disease diagnosis to the registration date) were reported. The median and the minimum and maximum of the Hoehn and Yahr (H&Y) stage were also described. To assess differences in the age of onset in our cohort compared to other populations (Abu Manneh et al., 2022; Ramchandra et al., 2025), we used a Chi-squared test to compare the number of patients with early-onset PD (EOPD; ≤49 years) and late-onset PD (LOPD; >50 years) according with the criteria reported by Riboldi et al. (2022) and Abu Manneh et al. (2022). To detect association between disease duration and disease severity, as indicated by the H&Y stage, a Spearman correlation was performed.

Moreover, the frequency of initial motor symptoms reported by the clinician, the location of the initial motor symptom, pharmacological treatment, and the years of delay in diagnosis among patients were compared using the Chi-squared test. Based on the understanding that pharmacological treatment changes as a function of age and disorder progression (Connolly & Lang, 2014), it was examined whether the pharmacological treatment in our population was associated with age (segmented into five groups: younger than 45, then divided every 10 years of age, and aged 76 and older), or disease severity (segmented into two groups, divided between those at stage 2 or less and those at stage 3 or more on the H&Y scale), by performing a Chi-squared test. For all analyses, results yielding a p-value of less than 0.05 were considered statistically significant.

All the statistical analyses were performed with the *R* (version 4.3.1) program.

### 2.5 DNA collection and Processing

Buccal swabs were collected from participants to obtain DNA. The procedure involved gently swabbing the inside of the cheek using sterile buccal swabs. Participants were instructed to refrain from eating, drinking, or chewing gum for at least 30 minutes before sample collection to minimize contamination. After swabbing, the swabs were carefully let to completely dry and placed into their envelopes. Envelopes were securely sealed and stored at 4° C until further processing.

The isolation of DNA was performed using the QIAamp DNA Mini Blood (Qiagen) following recommended procedure, and the concentration was measured using the Qubit ssDNA (Invitrogen) assay. Samples with a minimum DNA recovery of 2.1 μg were suspended in buffer solution and sent to the Lerner Research Institute at the Cleveland Clinic (Ohio, United States of America) as part of the collaboration with LARGE-PD. At this stage, the samples underwent a secondary assessment of concentration and a sex-check using a off-the-shelf TaqMan (Thermo Fisher Scientific) assay to ensure consistency. Subsequently, samples with a minimum concentration of 60 ng/μL and no sex mismatch were selected for genotyping.

### 2.6 Genotyping, Quality Control and Imputation

The selected samples were genotyped with the Illumina NeuroBooster Array, which contains ∼2M variants of which ∼90k are associated with 70 neurological conditions or traits (Bandres-Ciga et al. 2023).

Following genotyping, variant calling was performed using the Illumina Array Analysis platform. The Genotype Call (GTC) File was then converted to VCF format using the bcftools (Danecek et al. 2021) plugin GTC2VCF (available at https://github.com/freeseek/gtc2vcf).

To ensure variant quality, we cross-referenced the variants against the Illumina annotation files from both hg37 and hg38 genome builds. Variants that changed chromosomes during the liftOver process were excluded from the analysis. Additionally, we filtered out tri-allelic variants genotyped based on references other than the human genome reference, specifically those with alleles mapped to the ‘-’ strand. Further details can be consulted in https://github.com/MataLabCCF/callAndAnnotation

Genetic quality control was performed using the pipeline developed by the LARGE-PD consortium (available at https://github.com/MataLabCCF/GWASQC). Using Plink2.0 (*PLINK 2.0*, n.d.). Redundant variants, and variants and samples with missing data higher than 5% were removed. Variants were not filtered by Minor allele frequency (MAF). Additionally, we removed samples where the declared sex did not match the inferred sex using Plink1.90 (“PLINK 1.9,” n.d.; Chang et al. 2015) (F > 0.8 for males and F < 0.5 for females), samples with a heterozygosity rate of ±3 SD from the mean heterozygosity, and variants that failed the Hardy-Weinberg equilibrium (HWE) exact test (thresholds of 1e-6 for controls and 1e-10 for cases). After this, we calculated relationships using KING (Manichaikul et al. 2010) and generated a list of samples to be removed using NAToRA (kinship coefficient of 0.0884, second degree) (Leal et al. 2022).

All samples and variants that passed quality control were then imputed using the TOPMed Imputation server, which includes 133,597 reference samples and 445,600,184 genetic variants distributed across the 22 autosomes and the X chromosome. More information about the TOPMed Study (Taliun et al. 2021), the Imputation Server (Das et al. 2016), and Minimac Imputation (Fuchsberger, Abecasis, and Hinds 2015) can be found in their corresponding publications. Low-imputation-quality variants were filtered at a threshold of r2 < 0.3.

### 2.7 Ancestry estimation

Prior to ancestry estimation, PLINK files underwent linkage disequilibrium (LD) pruning to reduce SNP correlation using the following parameters: window size of 200 SNPs, step size of 50 SNPs, and an r^2 threshold of 0.2.

To perform the Principal Component Analysis (PCA), the data was merged with a subset of the 1000 Genomes (1KG) Project (Byrska-Bishop et al. 2022). The samples selected from the 1KG Project are non-admixed samples from African (AFR), East (EAS) and South Asian (SAS), European (EUR), and Native American (NAT) meta-populations (Shriner et al. 2023), making this dataset a reliable parental proxy reference dataset. Using the merged dataset, we pruned variants in linkage disequilibrium and performed the PCA using PLINK 2 (https://pubmed.ncbi.nlm.nih.gov/25722852/). To run the admixture analysis, we implemented ADMIXTURE 1.3 (https://dalexander.github.io/admixture/) using the supervised mode. Pie charts and histograms of the ancestry proportions and distribution were created using matplotlib in Python for PD cases and controls separately.

### 2.8 Variant Frequency Calculation and Association Analysis

A list of 141 variants associated with PD or parkinsonism risk, or PD-progression, was generated based on a curated list by Markopoulou et al. (Markopoulou et al. 2021) and selected top hits for available genetic studies of PD in Latin American or Mexican Individuals (Romero-Gutiérrez et al. 2021; Saffie-Awad et al. 2024; Loesch et al. 2021) were generated for a preliminary analysis.

The MAF of these selected variants for cases and controls was calculated using Plink1.9. Additionally, we performed an association analysis adjusting by age, sex and five principal components using Plink2.

### 2.9 Cross-Cohort Comparison

To enable comparison with the MEX-PD cohort sample, 11 variants with either nominal p-values in the adjusted association analysis or 0 frequency in the case or control group were selected and screened using the genotyping imputed data from the Global Parkinson’s Genetics Program (GP2; https://gp2.org/), in the Admixed American/Latin American (AMR), European (EUR), and African (AFR) ancestry groups. Variants were assessed using the same steps previously described. To facilitate comparison, we created bar plots for the MAF across cohorts employing the ggplot2 library in R version 4.1.2.

## 3. Results

### 3.1 Demographic data

Our study includes 531 subjects serving as controls (166 men and 365 women) and 470 participants with PD (272 men and 198 women). A higher proportion of men than women are represented within the PD group, while the opposite is observed in the control group (p<0.0001, Table 1). Regarding age and years of education, controls were younger and had more years of education; despite these differences being present between groups, they do not impact the variables analyzed to fulfill the aims of this report.

**Table 1.** Descriptive and lifestyle data of the sample. Significant results are marked in bold.

|  | Controls | PD patients | *p* |
| --- | --- | --- | --- |
| **n** | 531 | 470 |  |
| Men (%)<sup>a</sup> | 166 (31.3%) | 272 (57.9%) | **<0.001** |
| Women (%) | 365 (68.7%) | 198 (42.1%) |  |
| **Age<sup>b</sup>** | 58.37 ± 9.45 | 66.65 ± 10.31 | **<0.001** |
| **Years of education<sup>b</sup>** | 14.68 ± 5.22 | 12.39 ± 5.82 | **<0.001** |
| **Head injury or concussion<sup>c</sup>** |  |  |  |
| No | 403 (83.3%) | 328 (82.6%) | 0.870 |
| Yes | 81 (16.7%) | 69 (17.4%) |  |

<sup>a</sup>Group x Sex Chi square (2X2); significant differences were observed among all subgroups; *n* and percentage are reported.
<sup>b</sup>Student's t-test; Mean ± standard deviation is reported.
<sup>c</sup>Group x No/Yes Chi square (2x2); *n* and % is reported.

As a lifestyle factor associated with the onset of PD, the incidence of head trauma in controls and PD patients (only if it occurred before PD diagnosis) was compared. However, no significant differences were found between the control and PD patient groups (p>0.05; Table 1).

The geographical distribution of participants by state is illustrated in Figure 1. The majority of control participants coming from Mexico City, Querétaro, and Morelos. While, the states contributing with the most PD patients were Monterrey, Mexico City and San Luis Potosí.

### 3.2 Clinical data

The clinical characteristics of the patients are presented in Table 2. The average age at diagnosis was 59.9 years (SD 11.52), with a mean disease duration of 6.7 years (SD 5.9). Our sample was composed of 21.2% Early Onset Parkinson’s Disease (EOPD; <50 years) patients and 78.8% Late Onset Parkinson’s Disease (LOPD; ≥50 years) patients.

**Table 2.**
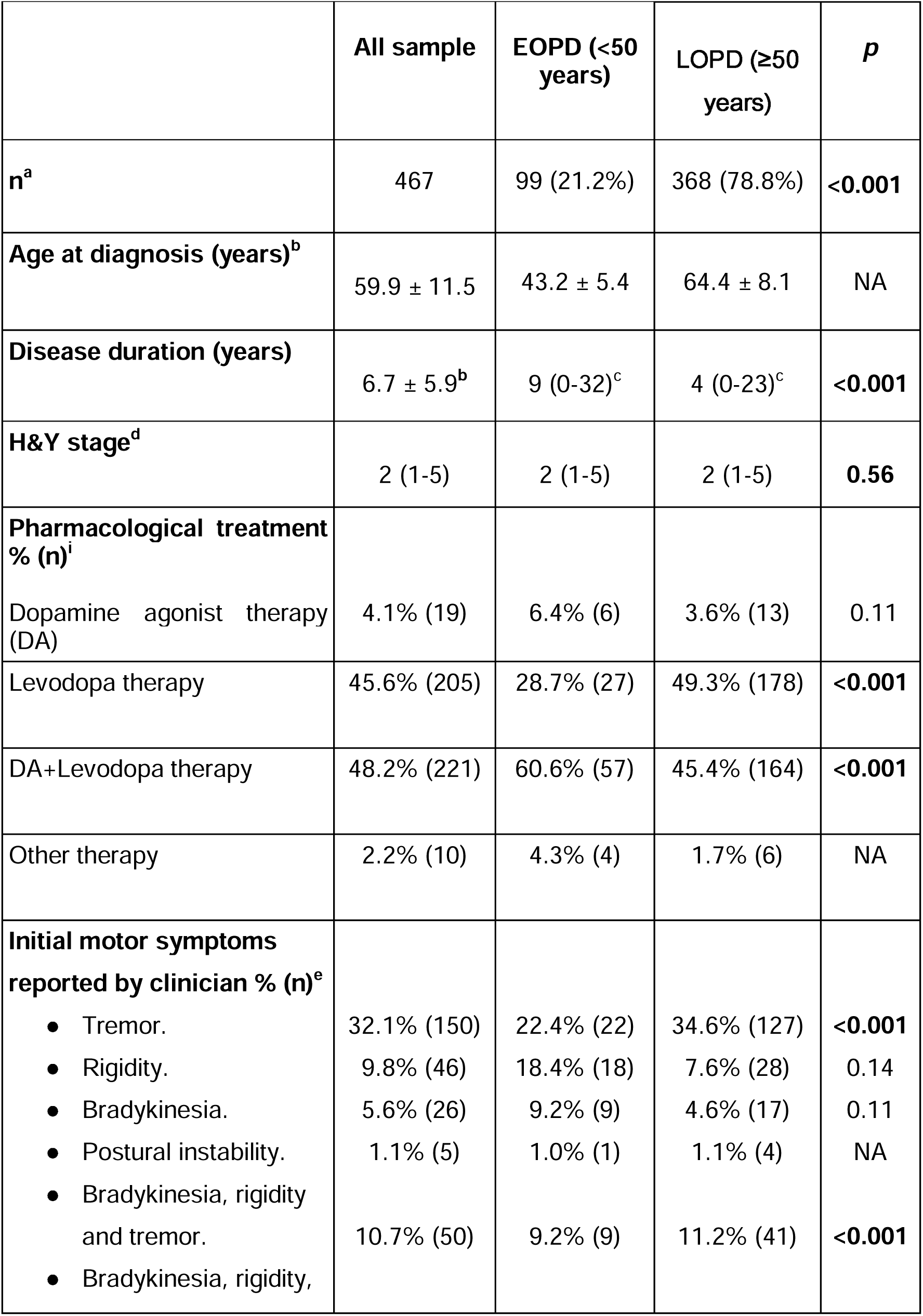
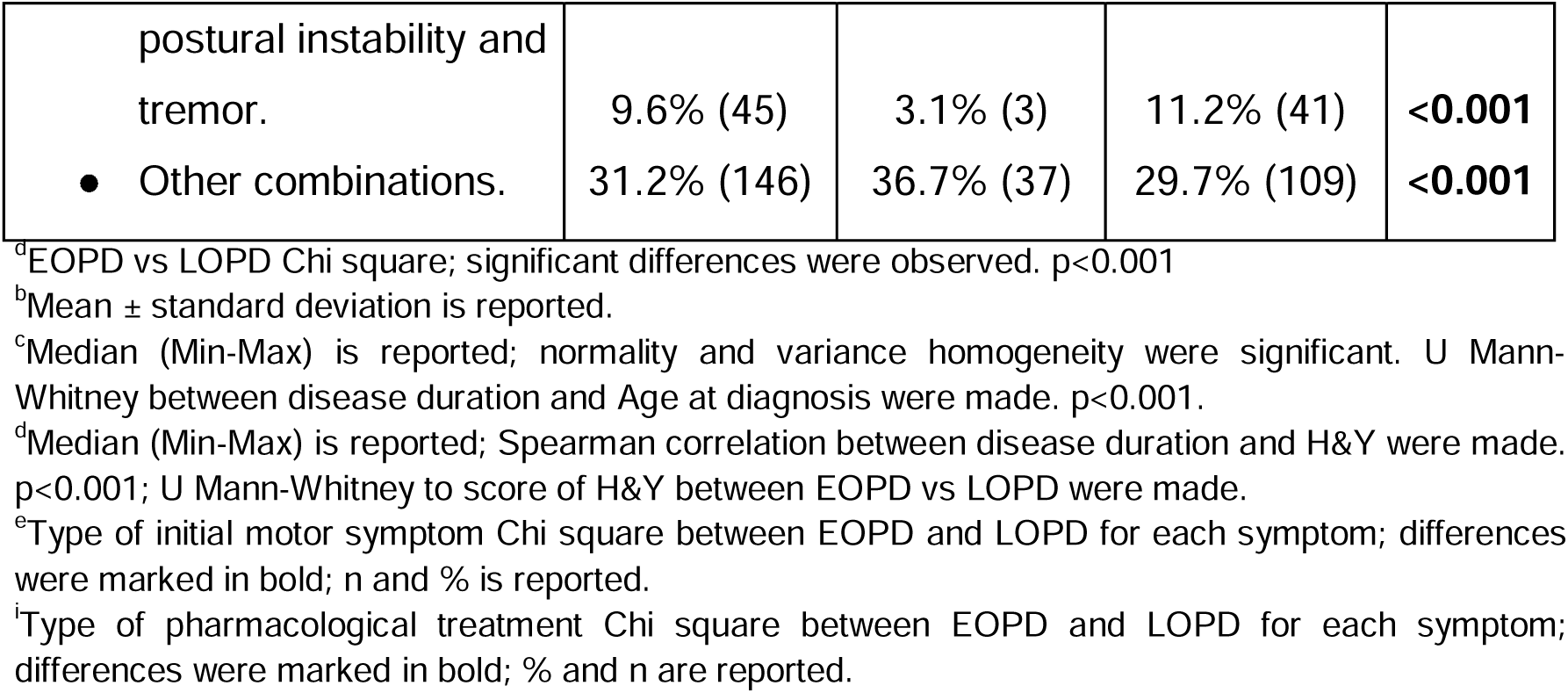
Clinical characteristics of the whole PD patient’s sample and the comparison as a function of the age of onset of PD. Significant results are marked in bold.

|  | All sample | EOPD (<50 years) | LOPD (≥50 years) | *p* |
| --- | --- | --- | --- | --- |
| **n<sup>a</sup>** | 467 | 99 (21.2%) | 368 (78.8%) | **<0.001** |
| **Age at diagnosis (years)<sup>b</sup>** | 59.9 ± 11.5 | 43.2 ± 5.4 | 64.4 ± 8.1 | NA |
| **Disease duration (years)** | 6.7 ± 5.9<sup>b</sup> | 9 (0-32)<sup>c</sup> | 4 (0-23)<sup>c</sup> | **<0.001** |
| **H&Y stage<sup>d</sup>** | 2 (1-5) | 2 (1-5) | 2 (1-5) | **0.56** |
| **Pharmacological treatment % (n)<sup>i</sup>** |  |  |  |  |
| Dopamine agonist therapy (DA) | 4.1% (19) | 6.4% (6) | 3.6% (13) | 0.11 |
| Levodopa therapy | 45.6% (205) | 28.7% (27) | 49.3% (178) | **<0.001** |
| DA+Levodopa therapy | 48.2% (221) | 60.6% (57) | 45.4% (164) | **<0.001** |
| Other therapy | 2.2% (10) | 4.3% (4) | 1.7% (6) | NA |
| **Initial motor symptoms reported by clinician % (n)<sup>e</sup>** |  |  |  |  |
| • Tremor. | 32.1% (150) | 22.4% (22) | 34.6% (127) | **<0.001** |
| • Rigidity. | 9.8% (46) | 18.4% (18) | 7.6% (28) | 0.14 |
| • Bradykinesia. | 5.6% (26) | 9.2% (9) | 4.6% (17) | 0.11 |
| • Postural instability. | 1.1% (5) | 1.0% (1) | 1.1% (4) | NA |
| • Bradykinesia, rigidity and tremor. | 10.7% (50) | 9.2% (9) | 11.2% (41) | **<0.001** |
| • Bradykinesia, rigidity, |  |  |  |  |
| postural instability and tremor. | 9.6% (45) | 3.1% (3) | 11.2% (41) | **<0.001** |
| • Other combinations. | 31.2% (146) | 36.7% (37) | 29.7% (109) | **<0.001** |

<sup>a</sup>EOPD vs LOPD Chi square; significant differences were observed. $p < 0.001$
<sup>b</sup>Mean ± standard deviation is reported.
<sup>c</sup>Median (Min-Max) is reported; normality and variance homogeneity were significant. U Mann-Whitney between disease duration and Age at diagnosis were made. $p < 0.001$ .
<sup>d</sup>Median (Min-Max) is reported; Spearman correlation between disease duration and H&Y were made. $p < 0.001$ ; U Mann-Whitney to score of H&Y between EOPD vs LOPD were made.
<sup>e</sup>Type of initial motor symptom Chi square between EOPD and LOPD for each symptom; differences were marked in bold; n and % is reported.
<sup>f</sup>Type of pharmacological treatment Chi square between EOPD and LOPD for each symptom; differences were marked in bold; % and n are reported.

At the time of their initial interview for this research, most patients were in a mild state of disease progression, as indicated by a median Hoehn & Yahr stage of 2 (min-max 1-5). A modest direct correlation was observed between disease duration and grade of disability (r= 0.33, p<0.001, r^2^=0.11).

Regarding pharmacological treatment, we found that 93.8% of PD patients were prescribed levodopa (either alone or in conjunction with a dopaminergic agonist [DA]), while 6.2% were undergoing other forms of pharmacological treatment (Table 2). There was an association between the most common pharmacological treatments in the sample (DA, DA+Levodopa and Levodopa alone) and etarian group (p=0.002; Figure 2.A). Additionally, we analyzed the association between the type of pharmacological treatment and disease severity, but no significant association was found (p= 0.65; Figure 2.B).

**Figure 2.**
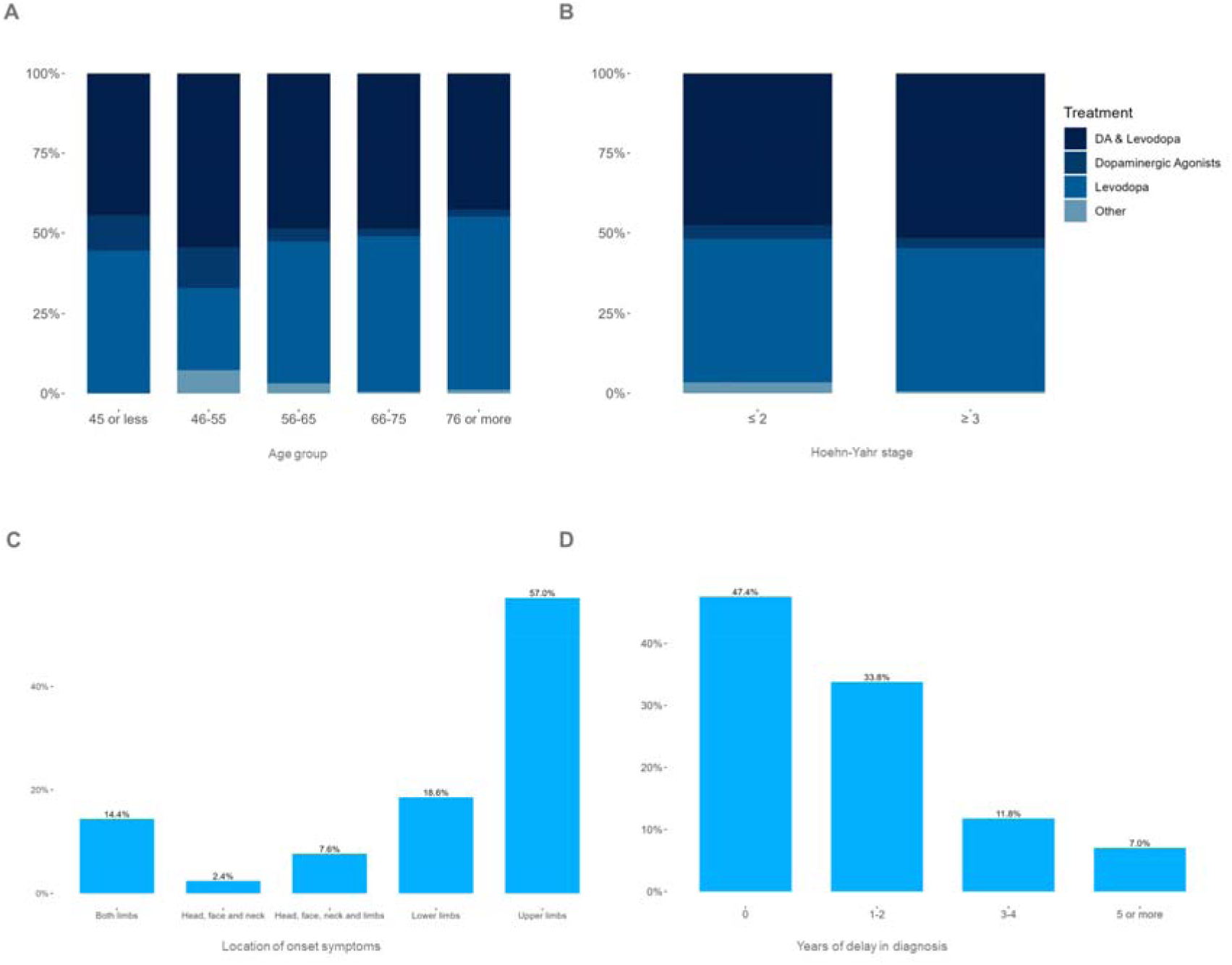
Treatment (Dopaminergic agonists, levodopa, and others) as a function of age group (A) and Hoehn & Yahr stage (B). There was a difference of treatment as a function of age (A; p=0.002) but not as a function of stage (B; p>0.05). Percentage of patients by the location of the initial symptoms (C). Years of delay in the diagnosis (D).

On the other hand, regarding the initial motor symptoms, tremor was the most common (32.1%, *p*<0.001; Table 2.), followed by the combination of differents motor symptoms; and the symptoms occurred mainly in the upper limbs (57%, *p* <0.001; Figure 2.C).

Finally, Figure 2.D describes the years of delay in the diagnosis of PD with respect to the time when the motor symptoms began. The majority (47.4%) of the patients were diagnosed the same year that the motor symptoms began (p<0.001), while 33.8% were diagnosed between the first and second year and 18.8% were diagnosed after three years or more.

### 3.3 Genetic data

#### DNA Collection and Processing

Until 2024, 695 genetic samples have been obtained meeting minimum DNA recovery of 2.1 ng of DNA. Of these 305 are from PD cases and 390 from controls. So far, 317 (113 cases, 204 controls) have been genotyped through Illumina NeuroBooster Array. Following quality control including the removal of redundant variants, variants with >5% missing data and failed HWE test, 1,580,839 variants remained.

After the exclusion of samples with >5% missing data, sex discrepancies, outlier heterozygosity and removal of related individuals, 294 samples were kept. Specifically, the removed samples included six for sex mismatch, four due to exceeding the missing data threshold (mind), four due to outlier heterozygosity, and nine, to relatedness.

These 294 samples and 1,580,839 variants were subjected to imputation using the TOPMed Imputation server. Following imputation and considering a r² < 0.3 threshold, 21,481,601 SNPs were retained for downstream analysis.

#### Ancestry estimation

Ancestry composition was estimated using ADMIXTURE in supervised mode, employing the participants of the 1000 Genomes Project as a reference population. This analysis revealed diverse parental contributions among the study participants. For PD cases, the highest proportion was found for EUR ancestry at 49.8%, followed by NAT ancestry at 46.1%. For controls, NAT ancestry was represented at 54.3%, while EUR ancestry accounted for 42.4%. In both groups, the sum of EAS and SAS ancestries comprised less than 1%. However, distribution of the ancestry contributions across the cohort revealed very similar patterns between the case and control groups (Figure 3).

**Figure 3.**
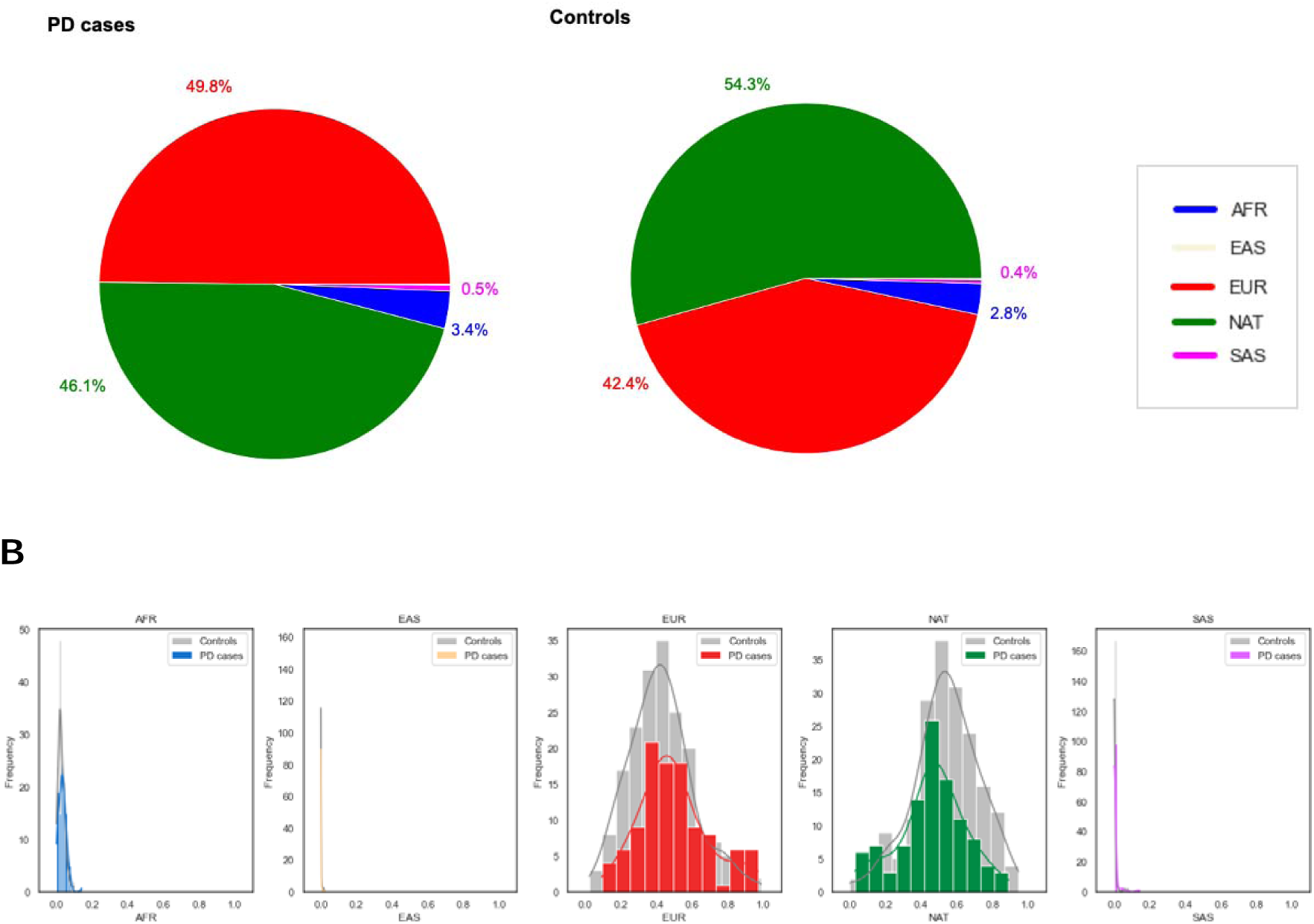
Ancestry composition in the MEX-PD cohort. A) Pie charts of the proportion of each ancestry: African (AFR), East Asian (EAS), European (EUR), Native American (NAT), and South Asian (SAS) in PD cases and Controls. B) Distribution of each ancestry in PD cases and Controls.

#### Allele Frequency Calculation and Cross-cohort Analysis

The allele frequency of 141 variants was calculated (Supplementary Table 2). Frequency of variants ranged between 0 and 50-55% for cases and controls, respectively. Of these, two variants (chr6:32218019:C:T and chr12:40340400:G:A) had a MAF of 0 either in the PD case or control groups respectively. These were selected to further screen in the GP2 AMR, EUR and AFR cohorts.

Furthermore, we analyzed these 141 variants using a generalized linear model (GLM) regression adjusted by age, sex and the first five principal components (Supplementary Table 3). Only nine variants had a nominal p-value (p < 0.05), and were further analyzed in the GP2 AMR cohort.

In summary, 11 variants were selected to compare the MAF and perform association tests in the GP2 cohort across AMR, AFR and EUR ancestry groups. The frequencies and results from association analyses are available in Supplementary Table 4 and 5, respectively.

Overall, the frequency of most of the SNPs in the MEX-PD cohort follows a similar pattern to the GP2-AMR group in the PD case vs control groups. However, we observed differences in the frequencies for chr3:122478045:C:T and chr6:71778059:C:T (Supplementary Figure 1)

**Supplementary Figure 1.**
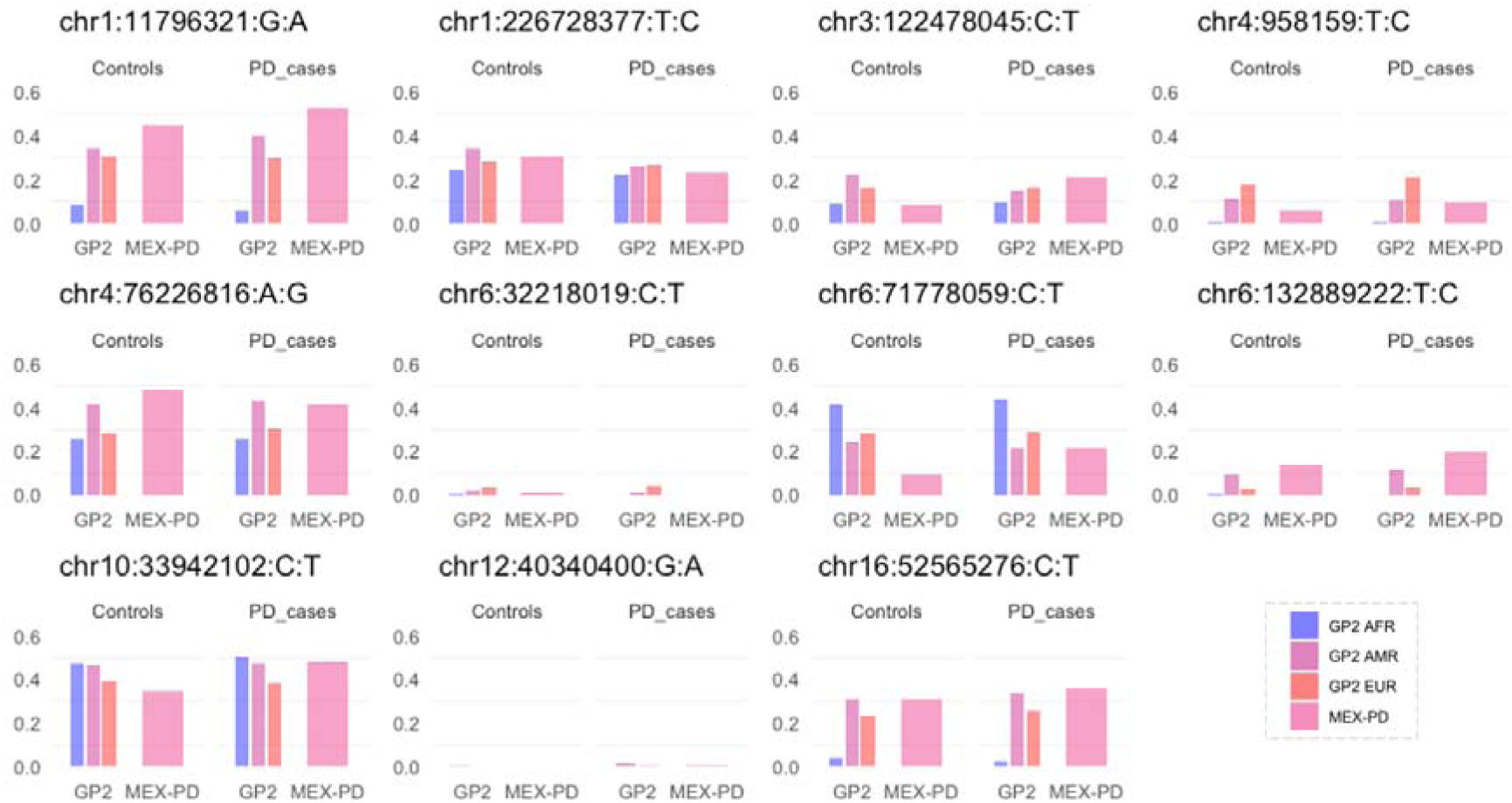
Frequency of 11 selected SNPs across the MEX-PD and GP2 cohorts. The three thin bars on the left correspond to the MAF in the African (AFR), Admixed American/Latin American (AMR), and European (EUR) ancestry groups from the GP2 cohort, while the thick bar on the right corresponds to the frequency of each SNP in the MEX-PD cohort.

As mentioned, nine SNPs were tested in the logistic adjusted model. Resulting odds ratios (OR) plus 95% confidence intervals of 6 out of the 9 SNPs were overlapping between GP2-AMR and MEX-PD cohorts, indicating a similar effect across studies. Furthermore, a nominally-significant protective effect was found in both datasets for the chr1:226728377:T:C variant.

In the case of chr6:132889222:T:C and chr6:71778059:C:T, there was only an overlap between confidence intervals, thus suggesting no statistically significant difference between them, but showing some uncertainty for true effect sizes. Finally, the odds ratio for chr3:122478045:C:T was different across cohorts, being protective (OR = 0.60) in the GP2-AMR cohort and of risk (OR = 2.31) in the MEX-PD cohort (Figure 4).

**Figure 4.**
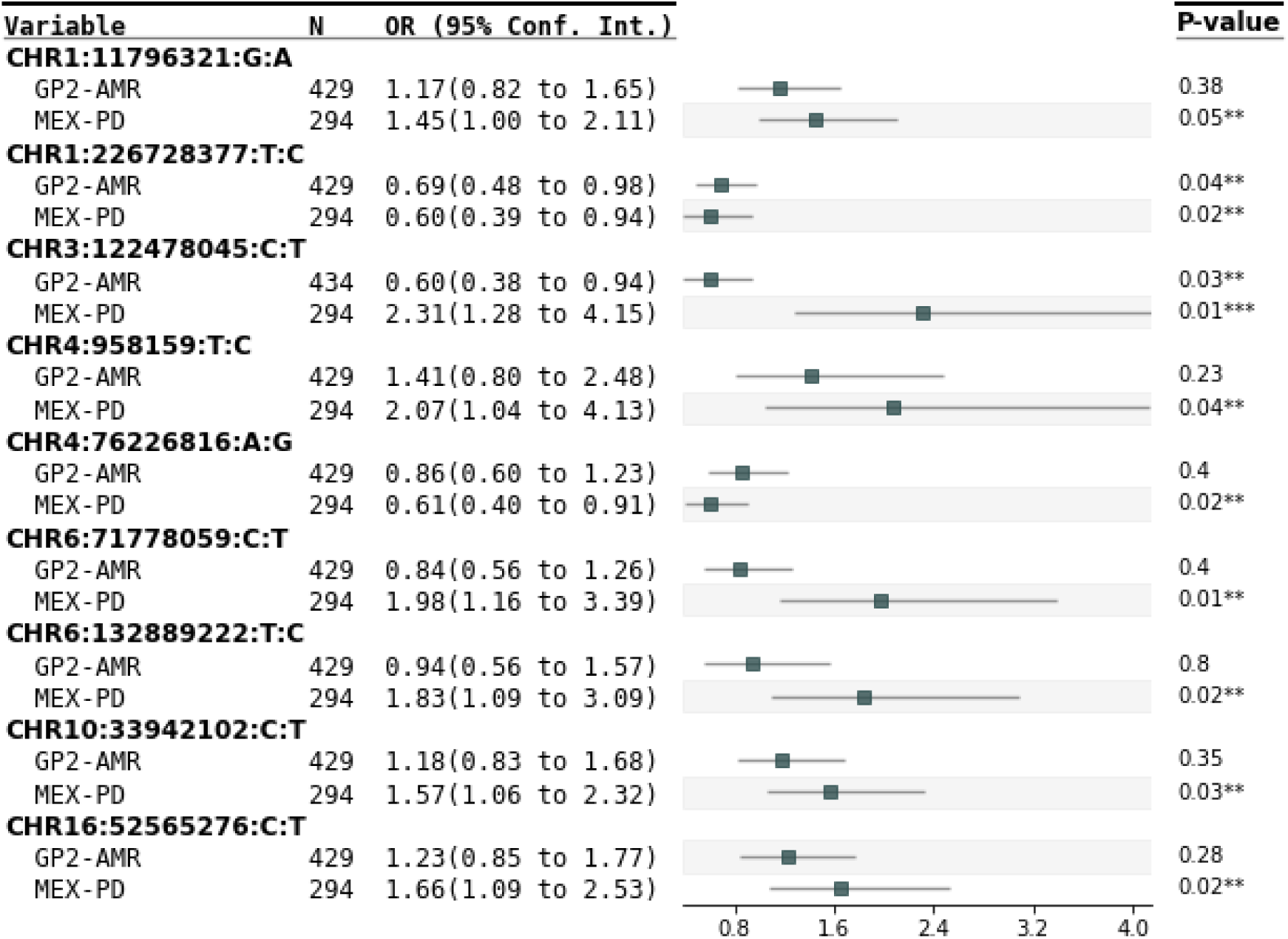
Forest plot showing OR estimates with a 95% confidence interval for 9 SNPs on Parkinson’s disease risk in the MEX-PD and GP2-AMR cohorts. P values of the association for each cohort are shown in the last column on the right and significance is marked with stars (*)

## 4. Discussion

MEX-PD is an effort to compile clinical and genetic data from PD patients and controls throughout Mexico, in order to highlight characteristics from Latino population and how they may impact the presence and progression of PD. We want to understand if the genetic admixture and the unique lifestyle conditions in Mexico might have a differential impact with respect to other populations.

Currently, we have data from 27 states (87% of the Mexico states) within the national territory, reaching almost all the 32 states in our country. Notwithstanding, in Mexico a systematic registry of PD patients (and other diseases) is lacking, thus MEX-PD seeks to register the largest number of diagnosed patients nationwide. The efforts of MEX-PD continue to reach this goal. As it was shown, in this Mexican cohort, the number of men was significantly higher (57.9%, Table 2) than women (42.1%) in the PD group, which is consistent with other populations (Tysnes & Storstein, 2017, Pringsheim et al., 2014). However, we have an opposite pattern for the control group: there are more women than men, thus, we will focus on registering more men in the next waves of the study. Regarding the dispersion of our records throughout the country, that most of them are located in the central part of Mexico, therefore, an effort will be made to register more patients and controls from the northwestern and south-east of Mexico in the future. Regarding this fact, there is a need to have more movement disorder specialists in every state of our country.

The age of onset of our sample was 59.9 years old (SD 11.52), which is consistent with most populations (Tysnes & Storstein, 2017, Khan et al., 2018). In a previous study reported by the National Institute of Neurology and Neurosurgery, located in Mexico City, the age of onset of PD was 57 years (Rodríguez-Violante et al., 2011). However, that report did not mention from which state of the country the patients came from. The difference in age of onset between studies could reveal how genetics or lifestyle factors, or their interaction, plays a role in the PD onset. Another potential factor that might explain the age of onset difference is the fact that the previous work obtained data only from the National Institute of Neurology and Neurosurgery, while the effort of MEX-PD is trying to incorporate data reaching all the states of the country.

On the other hand, evidence suggests that trauma is a risk factor for the appearance of PD (Gardner et al., 2018), although results are controversial (Goldam et al., 2018; Bower et al., 2003). In our cohort, the differences in head injury or concussion incidence between groups were not significant, suggesting head trauma is not a clear risk factor for PD in our population. However, this result should be taken as preliminary, expecting to assess this factor in a larger sample size to be collected in later waves.

Interestingly, 21.2% of the patients in our sample present early-onset Parkinson’s disease (EOPD). These preliminary findings suggest that the Mexican population may have a higher proportion of EOPD compared to European populations (5–15%; Bloem et al., 2021; Abu Manneh et al., 2022). In subsequent data collection phases, we plan to analyze whether specific genetic variants might be associated with age of onset in this cohort, using a larger sample to validate these observations.

Regarding the time elapsed between the appearance of the first symptoms and the diagnosis of PD (Figure 2.D), we can see that 47.4% were diagnosed the same year that the symptoms began, while 33.8% took between one and two years, and 18.8% took more than three years to be diagnosed. This information is relevant given that the longer the patient takes to be diagnosed, affects the estimates of the presence of the disease in the country population.

The pharmacological treatment that most of the patients received was levodopa (93.8%), followed by DA (52.3%). In fact, our data shows a trend that the use of DA decreases with age and levodopa increases, however the treatments were not associated with symptoms severity.

About motor symptoms, here it is documented that a third of patients (32.1%) had tremor as a first initial symptom, while a combination among tremor and other symptoms had the other third part 31.2%, and even this prevalence is maintained among EOPD and LOPD. Besides, these symptoms appeared mostly in the upper limbs. This initial symptomatology is consistent with what has been reported in other studies (Sveinbjornsdottir, 2016).

The present study represents the first report of a whole-genome genotyping approach in PD population in Mexico offering new insights into the genetic landscape of this disease within a previously understudied population.

After DNA collection and processing, rigorous quality control and imputation allowed us to retain a robust dataset of 294 samples and over 21 million SNPs for downstream analysis. Considering the important role that ancestry and admixture can play in explaining different traits (Horimoto et al. 2021; Moreno-Estrada et al. 2014), we performed an ancestry estimation using ADMIXTURE. The high proportions of EUR and NAT ancestries in both groups align with the known admixture patterns in Mexican people, where European and Native American contributions are predominant (Moreno-Estrada et al. 2014; Norris et al. 2018). The similar patterns of ancestry distribution between cases and controls suggest that the observed genetic differences cannot solely be explained by population stratification. However, local ancestry has been observed to modify disease susceptibility by influencing the effect of genetic variants in specific genomic regions (Beecham et al. 2022; Swart et al. 2021). Further analysis, such as admixture mapping or local ancestry analysis, is needed to disentangle these effects and better understand the genetic architecture of PD.

Furthermore, by carrying out allele frequency calculations and association analysis of 141 selected variants previously associated with PD we were able to present a general landscape of the MEX-PD sample while providing a guide for future studies.

Notably, chr6:32218019:C:T (rs8192591, located in the *NOTCH* gene) was present in controls but not in cases, and chr12:40340400:G:A (rs34637584, in *LRRK2*) was found only in cases, thus, highlighting their potential significance in the context of PD susceptibility. These findings are very similar to those from a study based on a Spanish population (Bandrés-Ciga et al. 2016), which aligns with genetic studies indicating that the European component in the Mexican gene pool primarily originates from Spain (Rubi-Castellanos et al. 2009)

Furthermore, in our cohort, nine variants were nominally significantly associated with PD risk. Remarkably, we observed a nominally significant risk effect (OR=1.45, p=0.05) of chr1:11796321:G:A (rs1801133, in *MTHFR*), matching with the previous findings of Romero-Gutiérrez et al (OR=2.02, p=0.043).

When the frequencies of 11 selected variants were contrasted to those from GP2 cohorts, most frequency patterns in the case and control groups from our cohort were comparable to the ones observed in the AMR population, except for chr3:122478045:C:T and chr6:71778059:C:T. This resemblance appears intuitive as the GP2 AMR cohort can be considered the closest population to Mexican (Vitale et al. 2024)

In light of this, using the GP2-AMR cohort, we examined the 9 nominally significant SNPs yielded in the association analysis from the MEX-PD cohort. Interestingly, only 2 of these 9 tested variants reached significance at a p <0.05 threshold in the GP2 AMR data. As the sample size for this dataset is about 1.5 times that of the current MEX-PD cohort, this might indicate that the remaining 7 variants could have a stronger or more specific association with PD in the MEX-PD cohort compared to the broader Latin American population.

In the cross-cohort comparison, no statistically significant differences were found between GP2-AMR and MEX-PD for 8 of the 9 variants. However, chr3:122478045:C (rs55961674, in *KPNA1*) showed a nominally significant association with PD in both cohorts, but in opposite directions. It was identified as protective in the GP2-AMR cohort, while it was associated with increased risk in the MEX-PD cohort. This SNP was one of the novel top hits from a PD GWAS meta-analysis, performed in cohorts of European ancestry only (Nalls et al. 2019) that associated rs55961674 to a modest risk (OR = 1.09)

Summarizing, most of the prioritized variables showed concordance between our cohort and the GP2-AMR data, thus suggesting that these variants may have consistent roles in PD across different Latin American populations and setting a valuable start point for further investigation. However, observed discrepancies could be attributed to population-specific genetic factors (Myles et al. 2008; Choudhury et al. 2014). Furthermore, we cannot know if the GP2-AMR is enriched for individuals from Latin American backgrounds that might differ from the Mexican population, such as those of Caribbean or South American descent. These findings highlight the complexity of genetic contributions to disease and the relevance of studying diverse populations.

Overall, our findings contribute to the growing body of knowledge on the genetic underpinnings of PD in Latin American populations, but particularly in the Mexican population. We encourage future studies to validate these results in larger, independent cohorts and to explore the functional consequences of the identified variants.

This work aims in the future to deepen our understanding of PD in the Mexican population, advocating for personalized treatments and improving patient outcomes in the Mexican population

### 4.1 Limitations

A limitation of this study is the lack of longitudinal follow-up of patients to evaluate the motor and non-motor symptoms progress. Moreover, our sample is highly concentrated at the center of the country, with a greater number of women in the control group and younger than the patients group. To manage this, we are making an effort to collect more information from the north and south-east of the country, as well as register older people and more men for the control group. Furthermore, regarding genetic analysis, our findings should be interpreted with caution as they are only preliminary and rather aim to prove the potential of the MEX-PD study, therefore necessitating validation in future stages of the study and in independent cohorts.

## 5. What is next for MEX-PD?

### 5.2 Neuroimaging

Patients and controls will be invited to attend the National Laboratory for Magnetic Resonance Imaging at UNAM campus Juriquilla in Queretaro city, to participate in an MRI session. This session will include *state-of-the-art* resting state functional imaging and high resolution multicontrast structural acquisitions in a 3T scanner, including T1, T2, (multishell) diffusion and neuromelanin weighted imaging. We expect to characterize the main functional and structural brain properties of patients with PD, aiming at identifying diagnostic and progression biomarkers for the disease and its non-motor symptoms.

### 5.3 Mental health and Cognitive analysis

The presence of mental health problems and cognitive impairment can affect the quality of life PD patients. As part of the evaluation of non-motor symptoms, the presence of anxiety and depression (state and trait) will be described. Also, global cognitive function will also be assessed to differentiate those with MCI. Moreover, we plan to evaluate different cognitive domains to find out which are the most affected in people with PD, and in this way future neuropsychological interventions will focus on these domains.

The results obtained from mental health and cognitive function will be compared between controls and patients, as well as their correlation with genetic data, structural and functional properties of the brain and environmental characterization.

### 5.4 Social community work

To engage with the community, we have established links with some Parkinson’s disease patient support groups through our Facebook page (https://www.facebook.com/RedMEXPD). We have organized webinars with neurologists and movement disorders specialists, published infographics related to PD and promoted training or events related to PD.

## Data Availability Statement

Metadata and QCed genomic data are available at upon request at https://gp2.org/research-outputs/gp2-required-agreement/

We downloaded the data from the 1000 Genomes 30x project, hosted at https://www.internationalgenome.org/data-portal/data-collection/30x-grch38. Subsequently, we extracted the samples based on the list provided by Shriner et al. (10.1016/j.xhgg.2023.100235). In this study, Shriner compiled a list of non-admixed samples using PCA and Admixture analyses for five meta-populations (African, European, East and South Asian, and Amerindian). The list is available upon request from Dr. Shriner.

Data used in the preparation of this article were obtained from the Global Parkinson’s Genetics Program (GP2; https://gp2.org). Specifically, we used Tier 2 data from GP2 release 7 [DOI: doi.org/10.5281/zenodo.10962119]. Tier 1 data can be accessed by completing a form on the Accelerating Medicines Partnership in Parkinson’s Disease (AMP®-PD) website (https://amp-pd.org/register-for-amp-pd). Tier 2 data access requires approval and a Data Use Agreement signed by your institution.

All code generated for this article, and the identifiers for all software programs and packages used, are available on GitHub (https://github.com/GP2code/MEX-PD-Latino) and were given a persistent identifier via Zenodo (https://zenodo.org/records/15185377)

## Acknowledgments

This work received support from Luis Aguilar, Alejandro León, and Jair García of the Laboratorio Nacional de Visualización Científica Avanzada. We thank Carina Uribe Díaz, Alejandra Castillo Carbajal and Christian Molina for their technical support.

We would like to thank the clinical neurologists who have contributed their valuable time and expertise for the recruitment and evaluation of participants: Sara Isaís Millán, Diana Deras, Lucero Ugalde, Omar Cárdenas, Ildefonso Rodríguez, Maritza Valadez, Moíses Rubio, Teresa Pérez. Those that contributed to collecting information and or sample processing from the participants: Roberto Estrella, Ingrid Salazar, Nelly Daniela López, María José Fernández, Alfonso Bravo, Diana Ramírez, E. Ivett Ortega-Mora, Sucel Iñiguez, Elissa López, Héctor Mederos, Abril Cerón, Brissa Peralta, Antinea Portillo, Itzel Hernández, Jonatan Serrano, Alan Morquecho, Jorge Aguilar, Diego Buoker, Itzel Ortiz, Diego Duarte, Karen Alberto, Dianalaura Hernández, Ana González, Gabriel de Anda, Luis Hernández, Francisco Ramírez, Sofía Hernández

We also thanks support from Mario Nandayapa Quartet, Yoloxochitl Sánchez-Guevara, Instituto de Biotecnología UNAM and Centro Cultural Manuel Gómez Morin, who facilitated recruitment of participants.

## Funding Sources for study

MEX-PD had the support from the American Parkinson’s Disease Association through a Diversity in Parkinson’s Disease Research Grant [APDA/D07]; CONAHCYT-FORDECYT-PRONACES grant no. [11311, 6390] and the support of the Global Parkinson’s Genetics Program GP2. GP2 is funded by the Aligning Science Across Parkinson’s (ASAP) initiative and implemented by The Michael J. Fox Foundation for Parkinson’s Research (https://gp2.org). For a complete list of GP2 members see https://gp2.org

ALF is a doctoral student of the *Programa de Doctorado en Psicología de la Universidad Nacional Autónoma de México* and she is a scholarship holder who received a grant scholarship [1222481] from CONAHCYT for her Psychology PhD studies.

PRP is a doctoral student of the *Programa de Doctorado en Ciencias Biomédicas de la Universidad Nacional Autónoma de México* and she is supported by the GP2 Trainee Network, which is part of the Global Parkinson’s Genetics Program and funded by the Aligning Science Across Parkinson’s (ASAP) initiative.

MER thanks fellowship support from the Rebecca L Cooper Medical Research Foundation [F20231230].

IFM and MIM are supported by the National Institutes of Health (NIH) [R01 1R01NS112499-01A1], ASAP-GP2 and the Parkinson’s Foundation.

Subaward LARGE-PD MJFF.

SA was supported by Programa de Apoyo a Proyectos de Investigación e Innovación Tecnológica–Universidad Nacional Autónoma de México (PAPIIT-UNAM) [IN208622].

AERC was supported by Programa de Apoyo a Proyectos de Investigación e Innovación Tecnológica–Universidad Nacional Autónoma de México (PAPIIT-UNAM) [IN217221].

AMR was supported by Programa de Apoyo a Proyectos de Investigación e Innovación Tecnológica–Universidad Nacional Autónoma de México (PAPIIT-UNAM) grant no. [IN218023].

## Financial Disclosures of all authors

The authors report no competing interests.

All authors have approved the final article.

